# Impact of Duration of Cessation of Mass BCG Vaccination Programs on COVID-19 Mortality

**DOI:** 10.1101/2020.08.20.20178889

**Authors:** Tareef Fadhil Raham

## Abstract

**Back ground:** BCG have heterogeneous immunity to certain pathogens other than *Mycobacterium tuberculosis* effect. At early times during COVID-19 pandemic heterogeneous immunity towards (SARS-CoV-2), was hypothesized and statistical correlation between of BCG vaccination practices and COVID-19 mortality variances among countries was statistically proved. These studies were criticized because of low evidence of such studies and possible confounding factors. For that reason, this study was designed to look for impact of duration of cessation of BCG programs on COVID-19 mortality looking for the hypotheses by different design and looking forward to support previous studies.

**Methods:** Total number of studied group is 14 countries which had stopped BCG vaccination programs.

Through applying stem-leaf plot for exploring data screening behavior concerning COVID-19Mortality for obsolescence duration of cessation of mass BCG vaccination programs, as well as (nonlinear regression of compound model) for predicted shape behavior for that group.

**Results:** Slope value shows highly significant effectiveness of obsolescence of cessation of mass BCG vaccination programs on COVID-19 mortality at P_-value_<0.000. Obsolescence of duration of cessation of mass BCG vaccination programs has strong negative association with COVID-19 mortality in countries which stopped BCG vaccination programs.

**Conclusiaon:** The longer the cessation duration of BCG programs, the higher the COVID-19mortality is, and vice versa.

## Introduction

Epidemiological and immunologic studies have shown reductions in morbidity and mortality following BCG immunizations in third world countries, suggesting that BCG may have some role in heterologous immunity to other pathogens^1^ furthermore, observational studies and randomized clinical trials show increase in childhood survival beyond the disease burden of the target disease ^1,2^

The biological substrate of these effects is mediated partly by heterologous effects on adaptive immunity, and on the potentiation of innate immune responses through ’trained immunity^3^. The term heterologous immunity refers to the immunity that can develop to one pathogen after a host had exposure to non-identical pathogens^4^

BCG vaccination significantly increases the secretion of pro-inflammatory cytokines, such as IL-1β and IL-6, which has been shown to play a vital role in antiviral immunity^5,6,7,8^. Furthermore, it has been proposed epigenetic reprogramming of immune cells, play an additional role through phenomenon conferring nonspecific immune memory to innate immune responses and termed ‘trained immunity3.

Many studies were done this year regarding statistical relation between BCG vaccination programs practicing and COVID-19 morbidity and mortality being less in countries carrying out such programs ^9,10^.

A rapid review^9^ and WHO brief ^11^ on 12 April 2020 evaluate the current evidence about the protective effects of BCG vaccine against acute respiratory infections and COVID-19. Although some have significant correlation brief states “WHO does not recommend BCG vaccination for the prevention of COVID-19” claiming that evidence is not strong enough yet.

Although heterogeneous immunity is not effective as homologous immunity to a specific pathogen which usually develops a very strong resistance to re-infection with the same pathogen ^12^,interests in heterogeneous protection is of great concern in SARS-COV-2 infection so far. Criticized studies hypothesized this theory and share the design of testing the BCG vaccination practices against morbidity and moratlity^9^. By testing the duration of cessation of BCG vaccination against COVID-19 mortality in this work, we are looking for the link by different design. The hypothesis in this study is the waning effect of vaccination as far as, the heterogeneous immunity waned with time^13^. So the primary aim of this study is supporting for previous studies through handling the question of immunity that’s BCG thought to be produced against (SARS-CoV-2) by different way that is the cessation duration of BCG programs in countries which stopped BCG vaccinations programs.

## Material and methods

Data regarding time of BCG cessation versus COVID-19 mortality is subjected to the test. Information on past BCG vaccination and cessation dates were collected from countries by data abstracted from published papers, reports, and available government policy documents retrieved through literature searches on PubMed and via the World Wide Web furthermore, we used immunization data available from the WHO-UNICEF estimates of BCG coverage site:

https://apps.who.int/immunization_monitoring/globalsummary/timeseries/tswucoveragebcg.html and WHO UNICEF review of national immunization coverage, 1980-2018 and immunization surveillance, assesment and monitoring data at site: https://www.who.int/immunization/monitoring_surveillance/data/afg.pdf

It was not appropriate or possible to involve patients or the public in this work. We used general practice level summated data and related publically published mortality statistics and national BCG protocols.

Total number of countries which stopped mass BCG vaccination programs is 14 countries. These countries are shown in table No. (1).

**Table No. (1):**
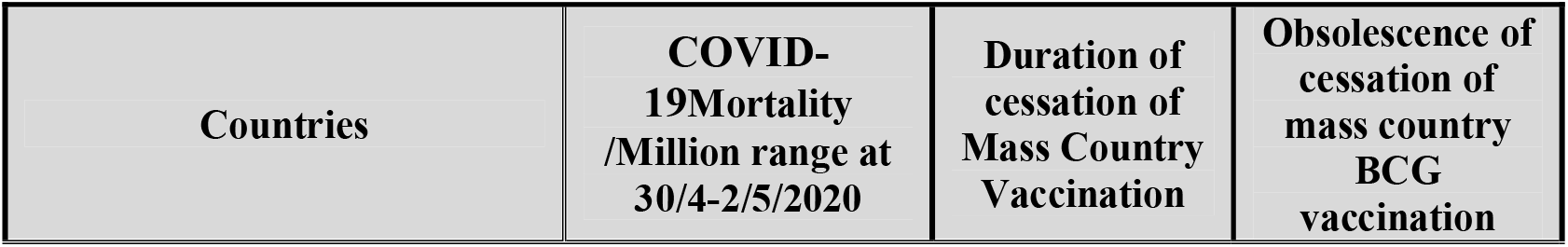
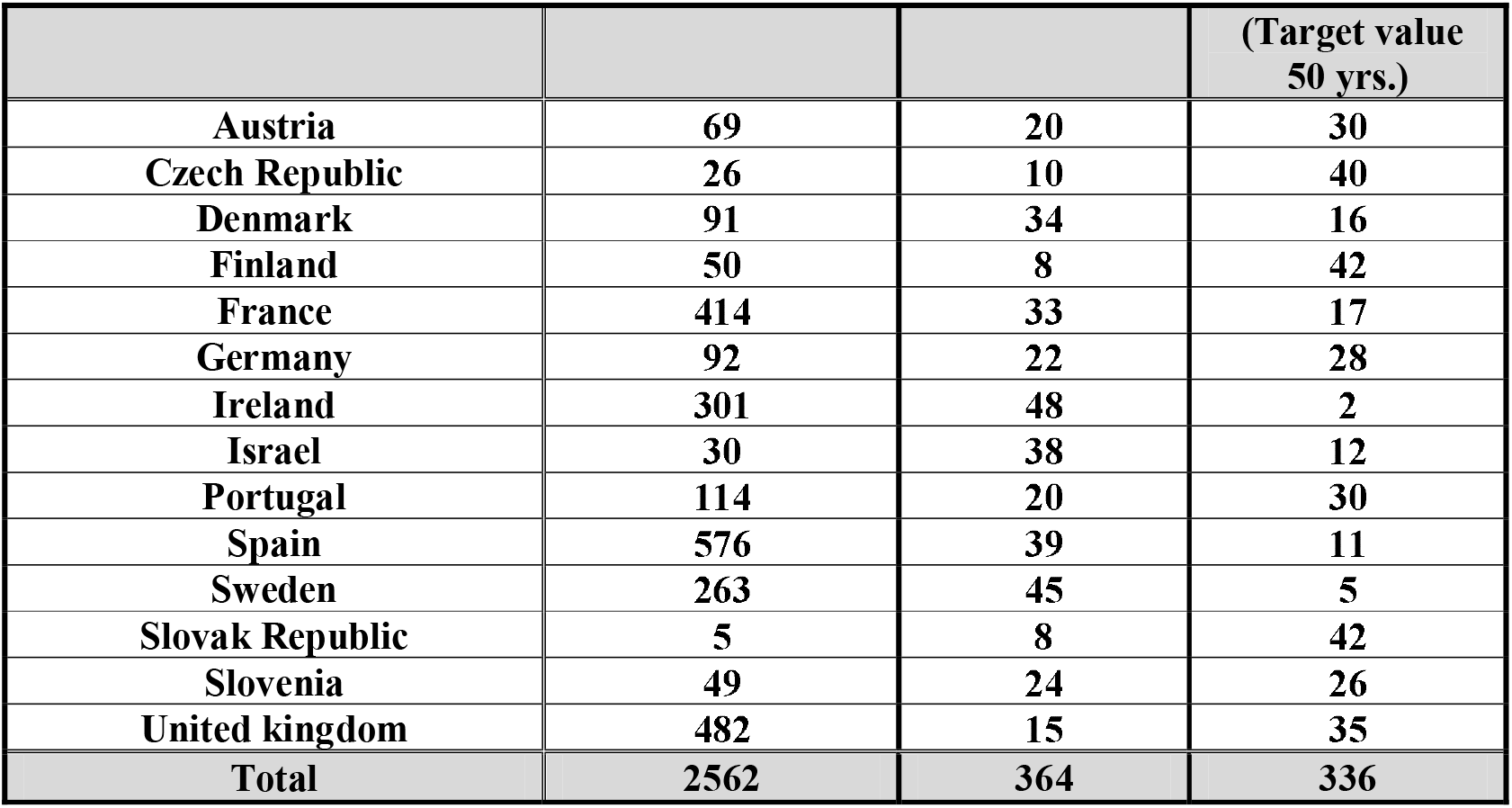
Duration of time since BCG cessation and obsolescence of cessation duration vs. COVID-19 mortality / million population inhabitant.

In addition to that, duration of obsolesce of cessation of mass BCG vaccination was calculated by subtracting cessation duration readings from the target value of (50) years which is choosed for statistical purposes providing that (no country has started BCG program in less than 50 years ago).

The reason of choosing the number (50) is the largest zero number for the closest longest period of time in Ireland country by rounding the period to the nearest zero numbers and then the time periods for each country were introduced to obtain the obsolescence duration for cessation of BCG vaccine for statistical purposes.

Furthermore, reasons for choosing obsolescence duration is to look for influence of vaccine on mortality from one side and for statistical reasons to bypass the problem of nonlinear models by conversion to a common logarithmic formula that cannot be achieved at zero when transforming to linear equation.

## Statistical Methods

The optimum model of highly fitted was checked among the several assumed models of non-linear regression, which can be transformed to linear equation, such as (logarithmic, inverse, quadratic, cubic, compound, power, s-shape, growth, exponential, and logistic), as well as the simple linear regression model was proposed also for predicted equations with their estimates such as (slope, intercept, correlation coefficient, determination coefficient, and regression ANOVA for testing the fitted model) are suggested for studying effectiveness of obsolescence of cessation of mass country BCG vaccination on COVID-19 mortality/million, and as illustrated in the table (2).

**Table No. (2):** Predicated equations for studying the effectiveness of obsolescence of cessation of mass BCG vaccination on COVID-19 mortality/million population inhabitant.

| Predicated Equations | Function |
| --- | --- |
| Model whose equation is $Y = b_0 + (b_1 * t)$. | Linear |
| Model whose equation is $Y = b_0 + (b_1 * \ln(t))$. | Logarithmic |
| Model whose equation is $Y = b_0 + (b_1 / t)$. | Inverse |
| Model whose equation is $Y = b_0 + (b_1 * t) + (b_2 * t^2)$. | Quadratic |
| Model defined by the equation $Y = b_0 + (b_1 * t) + (b_2 * t^{**2}) + (b_3 * t^{**3})$. | Cubic |
| Model whose equation is $Y = b_0 * (t^{**b_1})$ or $\text{Ln}(Y) = \text{Ln}(b_0) + (b_1 * \text{Ln}(t))$. | Compound |
| Model whose equation is $Y = b_0 * (b_1^{**t})$ or $\text{Ln}(Y) = \text{Ln}(b_0) + (\text{Ln}(b_1) * t)$. | Power |
| Model whose equation is $Y = e^{**}(b_0 + (b_1/t))$ or $\text{Ln}(Y) = b_0 + (b_1/t)$. | S-Shape |
| Model whose equation is $Y = 1 / (1/u + (b_0 * (b_1^{**t})))$ or $\text{Ln}(1/y-1/u) = \text{Ln}(b_0) + (\text{Ln}(b_1)*t)$ | Growth |
| Model whose equation is $Y = e^{**}(b_0 + (b_1 * t))$ or $\text{Ln}(Y) = b_0 + (b_1 * t)$. | Exponential |
| Model whose equation is $Y = b_0 * (e^{**}(b_1 * t))$ or $\text{Ln}(Y) = \text{Ln}(b_0) + (b_1 * t)$. | Logistic |

Shaded of Predicated Equations represents an optimal model.

In addition to that, stem-leaf plot was used for exploring data screening behavior concerning COVID-19 Mortality with obsolescence of cessation of mass BCG vaccination in countries previously given the BCG vaccine which are redistributed in three groups intervals.

All statistical operations were performed through using the ready-made statistical package SPSS, version 22.

## Results and Findings

Herein explore procedure known as (stem-leaf plot) has been proposed, since it produces summary statistics through graphical display. There are many reasons to do that, such as (outlier identification, description, and assumption checking). Data screening may show that presented unusual values, extreme values, gaps in the data, or other peculiarities. Exploring data can help to determine whether the statistical techniques that considers data analysis are appropriate.

The form 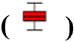 illustrated the core of actual behavior concerning studied data were included between the ordered observations which have less than two degree of standard values from the two sides, and the two edges of the box belongs to the first and the third quartiles and the median value fall between them. In addition to that, the observations that increased in more than three degree of standard values from the two sides would be assigned by a star and known as an outlier value.

Figure No. (1) represent graphically stem-leaf plot for studying behavior of effective distribution of obsolescence of cessation of mass BCG vaccination on COVID-19 mortality/million populations inhabitant.

**Figure No. (1):**
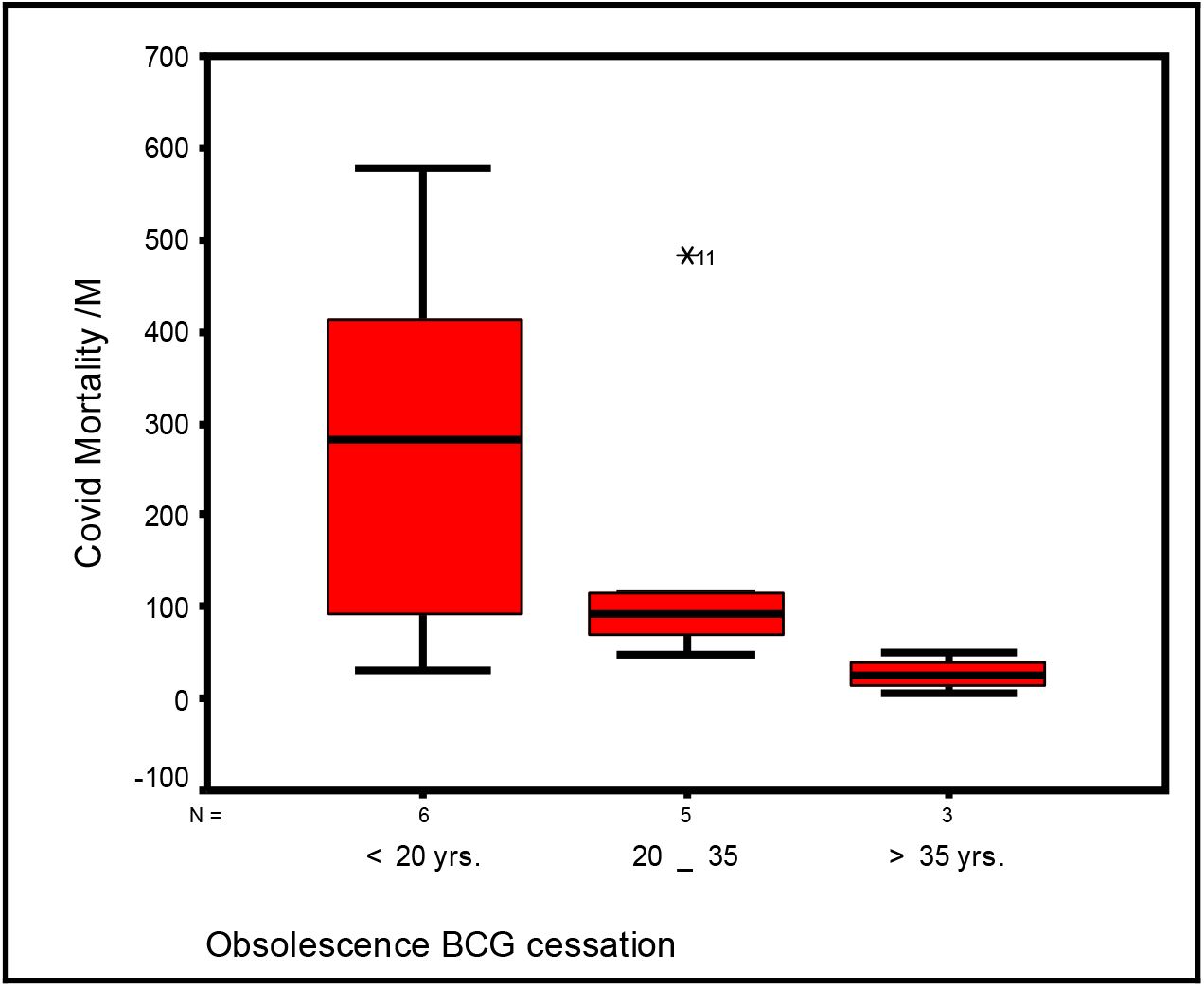
Stem-Leaf Plot of effectiveness obsolescence of cessation of mass BCG vaccination on COVID-19 mortality/million populations inhabitant.

It is clear from the redistribution in to the three intervals the effect BCG vaccine, effect of BCG vaccine being diminished or degenerated by stopping a given vaccine, and very importantly when the second interval of (20 – 35) years includes an outlier (United Kingdom), it did not affect the whole sample pattern of behavior or general mechanism for analyzing the effectiveness of the period of stopping the vaccine on Covid-19 mortality.

Table No. (3) and figure No. (2) show meaningful of nonlinear regression (compound model) tested in two tailed alternative statistical hypotheses. Slope value indicates that with increasing one unit of the obsolescence of cessation of mass BCG vaccination, there is negative effective on the unit of COVID-19 mortality/million., and estimated by (0.945442), which was recorded as highly significant effect at P_-value_<0.000, furthermore the relationship coefficient is accounted (0.57062) with meaningful and significant determination coefficient (R-Square = 32.56%). Other source of variations that are not included in studied model, i.e. “intercept” shows that non assignable factors which are not included in the regression equation, ought to be informative since about 369 cases would be an expected on Covid-19 mortality/million with the peak effectiveness of using vaccine at the earliest possible period, and rather than no significant P_-value_>0.05 was accounted, since probability level of significance simply stating not achieved.

**Table No. (3):** Effectiveness of Obsolescence of cessation of BCG mass vaccination on COVID-19 mortality/million

| Dependent variable Method... Compound -Shape Model "Covid -19 Mortality / M." |  |  |  |  |  |
| --- | --- | --- | --- | --- | --- |
| List wise Deletion of Missing Data |  |  |  |  |  |
| Simple Correlation Coefficient | 0.57062 (HS) | Meaningful Non Linear regression Tested in two tailed alternative Statistical hypothesis |  |  |  |
| R- Square | 0.32561 |  |  |  |  |
| F Statistic of Reg. ANOVA | 5.794 | Sign. F = | 0.0331 (S) <sup>(*)</sup> |  |  |
| Variables in the Equation |  |  |  |  |  |
| Variable | B | SE.B | Beta | t-test | Sig. of ( t ) <sup>(*)</sup> |
| Tb Prevalence / 100000 | 0.945442 | 0.02204 | 0.56517 | 42.904 | 0.0000 |
| (Constant) | 369.371 | 235.232 | - | 1.570 | 0.1423 |
| Predicted Equation - Logarithm-Shape Model |  |  |  |  |  |
| (Covid – 19 Mortality/M) = (369.371) * (Obsolesence Duration)<sup>0.945442</sup> |  |  |  |  |  |

<sup>(\*)</sup> HS: Highly Sig. at P<0.01; S: Sig. at P<0.05

Figure No. (1): Long term trend of scatter diagram effectiveness Obsolescence BCG cessation on COVID-19 mortality

**Figure No. (2):**
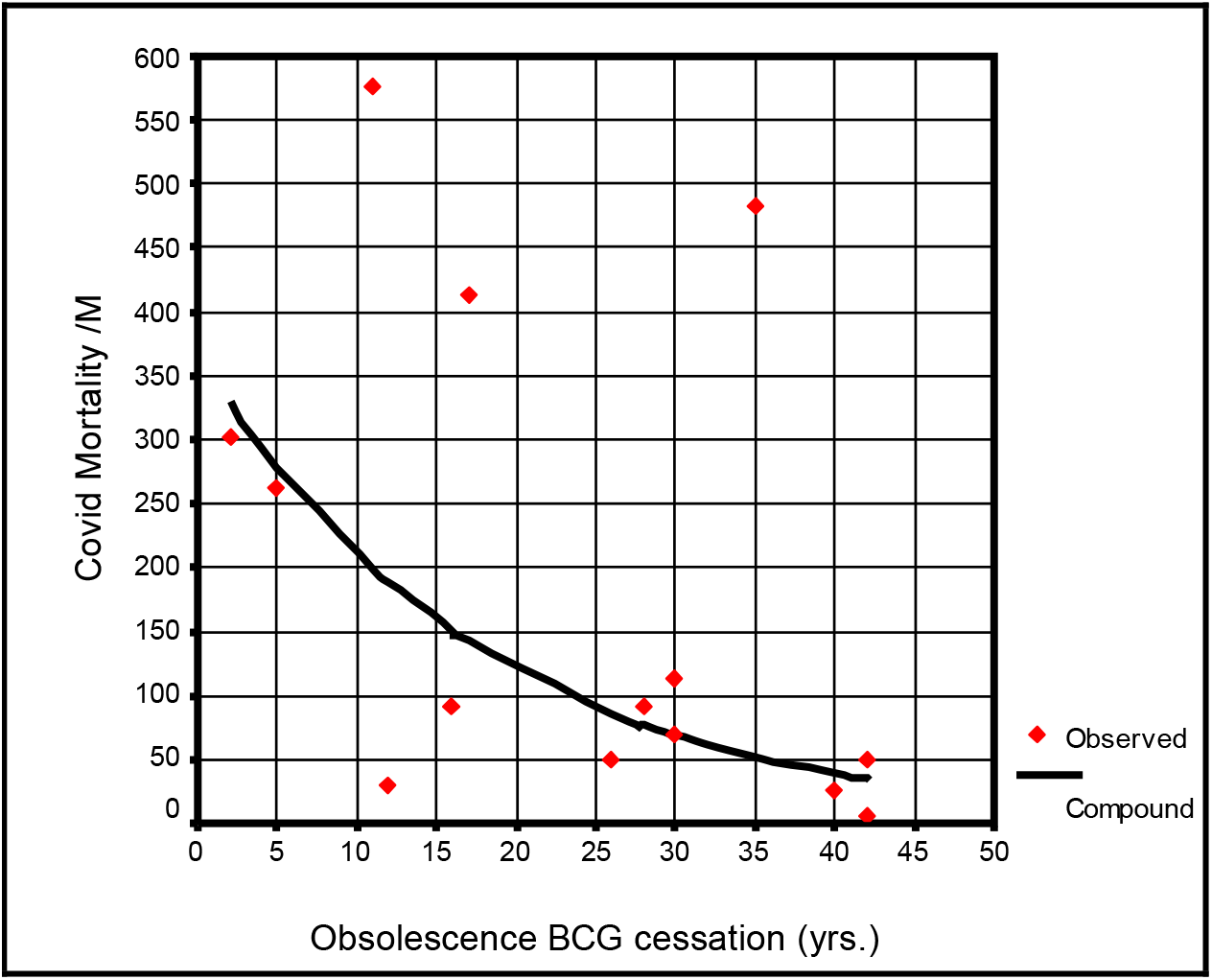
shows the long term trend of scatter diagram effectiveness of obsolescence of cessation of mass BCG on COVID-19 mortality/million.

## Discussion

Results reflect statistical correlation between BCG duration of obsolesce of cessation (when cessation time is diminished or degenerated) and COVID-19 mortality and being strongly negatively associated (Fig.1, Fig.2, table3). This association reflects the shorter cessation period of BCG vaccination the lesser COVID-19 mortality and vice versa.

Looking for duration of cessation of BCG vaccination program as factor is a new idea up to my knowledge to be tested in relation to COVID-19 mortality supporting the hypotheses of effectiveness of BCG vaccinations programs in prevention of COVID-19 mortality.

Previous studies regarding BCG effectiveness where criticized by possibility of significant bias from many confounders. Possible confounders: differences in national demographics and disease burden, testing rates for COVID-19 virus infections, and the stage of the pandemic in each country9.

In this study although above confounders cannot excluded but the design of the study is different from other studies. The influence of time proved in this study is of concern since previously criticized studies prove significant associations results with practices, results of this study and previous studies consolidate each other through different study designs.

In one study done before there was a positive significant correlation (ρ=0.54, p=0.02, linear correlation) between the year of the establishment of universal BCG vaccination and the mortality rate. This study did not assess the impact of cessation but the time of establishment of vaccination programs and includes both groups of countries which continue and not continue the vaccination programs, although the design is different again her, but similar finding consolidates evidence to each other^10^.

Another more recent study shows highly significant negative association between prevalence of TB and COVID-19 mortality which furtherly support such hypothesis regarding heterogeneous immunity of Mycobacterium through latent TB infections which its prevalence is proportional to TB prevalence^14^.

Highly significant association in this study support the hypotheses already raised about beneficial effect of vaccination and hence impact of its cessation.

The theoretical background for this association is protection role of heterogeneous immunity produced by BCG vaccine. As far as the duration of cessation of vaccination in this study have impact on mortality, its results might reflect the waning effect of heterogeneous immunity but still this finding should be confirmed by control clinical studies and immunological studies. For such clinical studies we need more time to wait to see such studies and this give importance to do such studies in this time. While this study states that duration of cessation of BCG programs have impact on COVID-19 mortality / million populations inhabitant explains more possible factor in variances in COVID-19 mortality through different countries in one hand and strengthen the evidence for early intervention at this critical time in another hand.

## Conclusion

Duration of cessation of BCG vaccination have significant effect on mortality due to COVID-19 and this study supports the evidence of effect of BCG vaccination in prevention of COVID-19 mortality.

## Data Availability

Data used are publicly available

## Recommendations

Decisions on BCG vaccination program cessation might be reconsidered.

## Ethics and dissemination

Ethical permission is not necessary as this study analyzed publically published data and patients were not involved.

There is no conflict of interest.

There is no funding received.

## Acknowledgment

I am deeply grateful to Emeritus Professor Abdulkhaleq Abduljabbar Ali Ghalib Al-Naqeeb, Ph.D. in the Philosophy of Statistical Sciences at the Medical & Health Technology college, Baghdad-Iraq, for his assistance and support in data analysis, interpretations of finding results, and revising the paper.

